# Recombination shapes 2022 monkeypox outbreak

**DOI:** 10.1101/2022.08.09.22278589

**Authors:** Ting-Yu Yeh, Zih-Yu Hsieh, Michael C. Feehley, Patrick J. Feehley, Gregory P. Contreras, Ying-Chieh Su, Shang-Lin Hsieh, Dylan A. Lewis

**Affiliations:** Auxergen Inc., Columbus Center, Baltimore, MD 21202, USA; Lake Washington High School, Kirkland, WA 98033, USA; Department of Thoracic Surgery, Chi-Mei Medical Center, Tainan City 710, Taiwan; Department of Biotechnology and Bioindustry Sciences, National Cheng Kung University, Tainan City 710, Taiwan; MacKay Memorial Hospital, Taipei City 104, Taiwan; Monte Vista High School, Danville, California 94526, USA

**Keywords:** Monkeypox, tandem repeats, polymorphism, site-frequency spectrum, linkage disequilibrium, recombination

## Abstract

**Background:** On July 23, 2022, the WHO declared monkeypox outbreak a global health emergency. Here we analyze monkeypox virus (MPXV) sequences during 2022 pandemic to investigate whether the virus is adapting for better survival and transmission among the human population.

**Methods:** By studying tandem repeats (TRs) and linkage disequilibrium (LD), we analyzed 415 MPXV sequences from January 1 to July 20, 2022 worldwide.

**Finding:** The 2022 MPXV population has diverged into 4 lineages and 11 subgroups based on various TRs and their copy numbers. LD analysis also shows that virus has evolved into 3 new lineages. We identify 8 new recombinants (six from Slovenia, one from Australia, one from Italy) using TR analysis and 3 recombinants (two from Germany, one from Spain) using LD analysis.

**Conclusion:** Our results indicate that the MPXV genome is evolving and expanding quickly during the 2022 pandemic. We conclude that in combination with genomic surveillance, TR analysis, as well as LD analysis, are useful tools with which to monitor and track phylogenetic dynamics and recombination of monkeypox transmission.

## Introduction

The 2022 monkeypox outbreak represents the first time this disease has spread widely beyond Central and West Africa. Initially identified in the United Kingdom in May 2022, monkeypox case numbers quickly increased in Europe, North and South America, Asia, Africa, and Oceania. On July 23, 2022, the WHO declared monkeypox outbreak a global health emergency. On August 12, 2022, a total of 35,032 cases were confirmed in nearly 80 countries (<u>Monkeypox data explorer.</u> *Our World in Data*).

Most monkeypox virus (MPXV) sequences from the 2022 outbreak belong to the B.1 clade (except two cases from the United States). In this study, we analyze MPXV sequences during 2022 pandemic to investigate whether the virus is adapting for better survival and transmission among the human population.

In a rapidly evolving poxvirus, adaptation is simultaneously driven by two mechanisms: recombination (gene copy number variation, fast) and single nucleotide variants (SNVs, slow) at the same loci (Sasani, et al., 2018). Recombination generates new phenotypes with greatly altered disease potential which are better suited to viral survival. It has been shown that vaccinia viral DNA is swapped back and forth ∼18 times per genome in a single round of infection (Qin and Evans, 2014) to make recombinant phenotypes. Recombination in poxvirus genomes has been commonly detected by selection or screening in laboratory animals or cell culture for more than 60 years (Fenner & Comben, 1958). Gershon et al. described recombination of four capripoxvirus isolates during natural virus transmission by analyzing physical maps of the viral genome (Gershon et al., 1989). However, very little is known about poxvirus recombination in nature due to its relatively large genome size and lack of genomic surveillance data, which is now becoming available.

Our efforts focused on discovering the variability of the MPXV genome via recombination to determine the potential risk of new viral strains. Tandem repeats (TRs) were first identified within the inverted terminal repeat of vaccinia virus DNA with a 70-base-pair sequence arranged in two blocks of 13 and 17 copies, respectively (Wittek and Moss, 1982). Other poxvirus TRs are small pieces of DNA sequences (3-25 nucleotides), and their sequences and copy numbers vary among different poxvirus family members or isolates (Kugelman et al., 2014). The insertions and deletions of TRs are common events among these poxviruses. (Coulson and Upton, 2011). Therefore, these TRs exhibit high rates of variation, and they represent a target for poxvirus gene truncation and variation (Kugalman et al., 2014; Hatcher et al., 2015). Here we report the first evidence of natural recombination of monkeypox virus by analyzing TRs. We also use linkage disequilibrium, a well-known SNVs-based analysis, to detect new lineages and recombination in MPXV genome of 2022 pandemics.

## Results

### Tandem repeats (TRs) as novel genetic markers of monkeypox virus to detect new lineages

To determine the genomic diversity in monkeypox genomes in the 2022 outbreak, we first searched TRs in 415 available MPXV sequences (B.1 clade) worldwide from January 1 to July 20, using the Tandem Repeat Finder algorithm (Benson, 1999). The advantage of this method is that it eliminates the bias caused by sequence alignment error, especially in low complexity sequences like TRs. Based on the criteria of the alignment score (>100) and length (>7 base pair, bp), we identified 6 TRs with variations in their copy numbers (Figure 1A and 1B). TR A/E have identical 16 bp sequences of inverted repeats (5’-TAACTCTAACTTATGACT-3’ and 5’-AGTCATAAGTTAGTTA-3’) at both ends of MPXV genome (Figure 1B). The viral populations were further categorized into six groups based on TR numbers (TRNs) of TRA/E (Figure 1C). 378 cases (90.6%) were associated with TRN=7.9 versus 14 cases of TRN=15.9 (3.4%), which were collected in the USA (ON959133, ON959134, ON959135, ON959136, ON954773, ON959131, ON959132), Belgium (ONON622712, ON622713, ON880419, ON880420, ON880421, ON880422) and the Czech Republic (ON983168). One case of TRN=5.6, TRN=3.6, and TRN=2.6 was found in United Kingdom (ON 619837, ON619835, ON022171). There were 21 cases with different TR numbers between TR A and E (“mismatch” in Figure 1C). This result shows that genome diversity can be grouped by TR polymorphism among MPXV populations in the 2022 pandemic.

**Figure 1.**
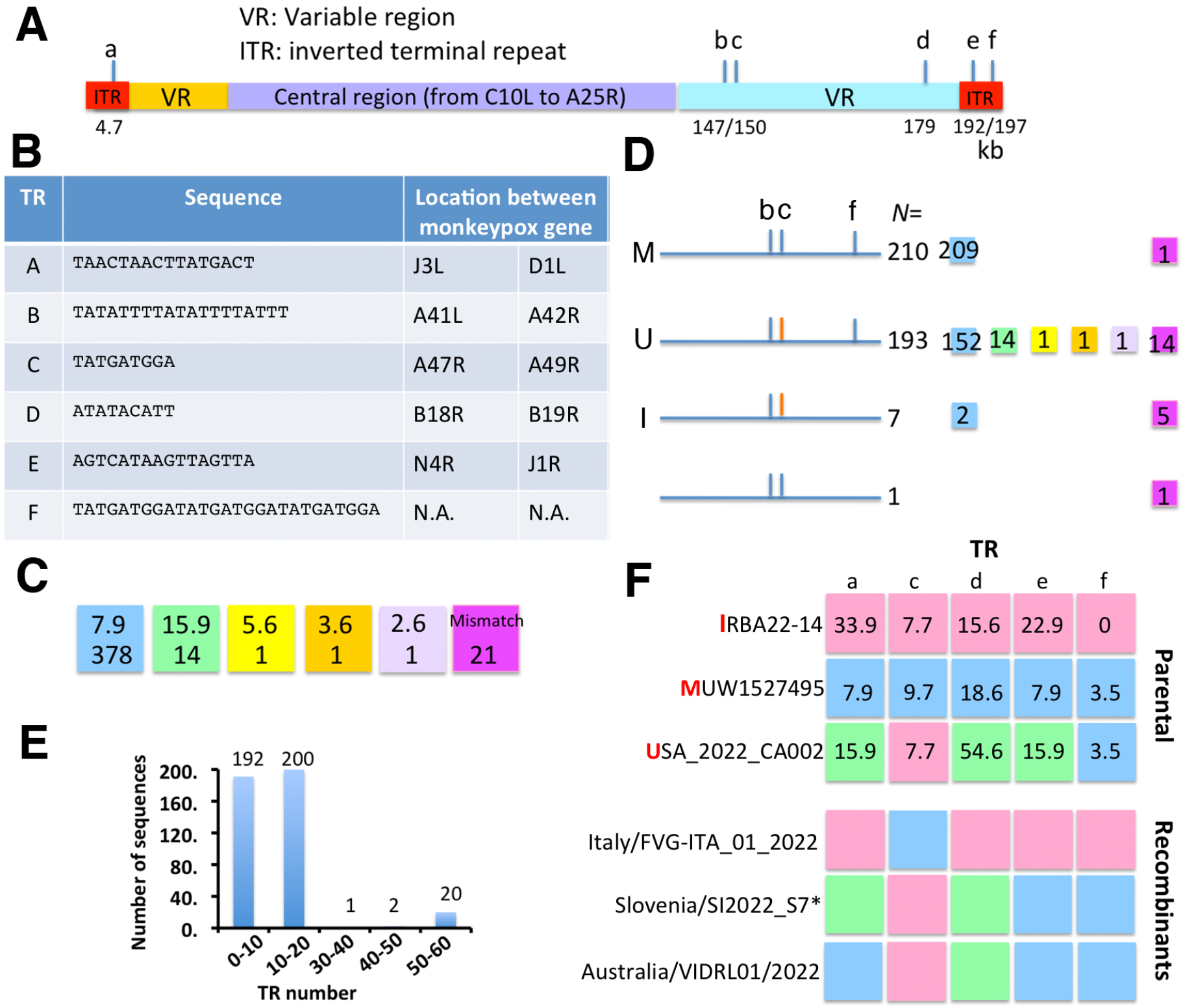
TR analysis of genetic diversity and recombination of MPXV genome in 2022 pandemics. (A to E) Characterization of tandem repeats (TRs) of MPXV genome in 2022 pandemics. (A) The map of MPXV genome. The location of TR A to F (a-f) are indicated. (B) Summary of TR sequences and locations in this study. (C) The sequence variation of TR A/E among MPXV sequences (B.1 clade) from January 1 to July 20, 2022 (*N*=415). Upper: TR numbers (TRNs), bottom: case numbers. “Mismatch” indicates that TRNs are different between TR A and E. (D) Sequence variation was represented by TR C/F and TRA/E. TRN=9.7 and TRN=7.7 of TR C are labeled as blue and red line, respectively. Group M, U, and I are named based on the representative sequences: MUW1527495 (M, ON019276) or USA_2022_CA002 (U, ON954773), and IRBA22-14 (I, ON755040). (E) Distribution of TR D based on TR numbers. (F) Graphical representation of recombination based on TR polymorphism. Each box (a, c, d, e, f) contains individual TRNs of TR A, C, D, E, F with different colors associated with the parental viral sequences: IRBA22-14 (I, ON755040), MUW1527495 (M, ON019276) or USA_2022_CA002 (U, ON954773). Here “parental” only refers to their phylogenic relationship among the sequences, not to the routes of transmission. Recombination is defined when a new virus gains a TR with different TRNs, which leads to the mosaic pattern of the genome. Asterisks include six Slovenia cases that may have evolved into a new lineage.

TR B, C, D, and F are direct repeats and located at either the intergenic regions (TR B, C, D) or 3’ inverted terminal repeat (TR F) (Figure 1C, detail information of TR B, C, D in Supplementary Figure 2-4). Each TR C and F contain one and three copies of 9 bp sequence (5’-TATGATGGA-3’), respectively. While the majority of viral sequences contain TR F (TRN=3.5, 97.1%), 50.8% and 48.2% of total samples either have TR C with TRN=9.7 and TRN=7.7, respectively (Figure 1D). Based on TR C/F pattern, the viral populations can be classified into 4 lineages (M, 210 cases; U, 193 cases; I, 7 cases; and one uncategorized, Figure 1D). Further, in combination with TRNs of TR C/F and TR A/E, we are able to categorize viral populations into 11 subgroups. TR D is a 9 base-pair sequence (5’-ATATCATT-3’) with more various copy numbers, ranging from TRN=2 to 54.6 (Figure 1E). Interestingly, the sequences with high TRNs of TR D (TRN>30) also contain higher TRNs of TR A/E (TRN>15.9, 20 cases) in the group U, indicating that lineage U has more TR diversity than the others.

Taken together, our data demonstrate that TRs diverged frequently during natural transmission within the B.1 clade and that the virus is evolving as its population expands (Zeng’s *E* = -1.65, Achaz’s *Y*=-2.52, *p*< 0.001).

### TR analysis identifies recombination of monkeypox genome in natural transmission

Poxvirus recombination between two co-infecting parental viruses generates genetic diversity (Ball, 1987; Evans et al., 1988; Sasani, et al, 2018). MPXV recombination in natural infection has not been reported to date. Using TR polymorphism, we identified 8 genomes with recombinant crossovers (Figure 1F). Case FVG-ITA-01 (ON755039) in Italy may be generated from parental sequences from the group I and M. Case VIDRL01 (ON631963) in Australia comes from parental sequence of the group M and U (Figure 1D), as well as six cases of Slovenia cases (ON838178, ON631241, ON609725, ON754985, ON754986, ON754987). This is the first report of recombination of MPXV in natural transmission to our knowledge. Our results also suggests that six Slovenia cases may have evolved into a new lineage.

### Linkage disequilibrium (LD) analysis shows new lineages and recombination

We then employed single nucleotide polymorphism (SNPs) analysis using the DNASP v6 algorithm to detect the occurrence of recombination (Rozas’s Za=0.0005, *p*<0.05) (Rozas et al., 2011). We also used Haploview algorithm to visualize the patterns of linkage disequilibrium (LD) between variants with minor alleles in at least two MPXV isolates and to detect the possible recombination (Figure 2A). In the absence of evolutionary forces or natural selection, *D*’, the normalized coefficient of LD, converges to zero along the time axis at a rate depending on the magnitude of the recombination rate between the two loci. Since most SNPs were at very low frequencies, many SNP pairs had low values of squared coefficient of correlation (*r*^2^) and the log of the odds (LOD) (Fig. 2D and 2E).

The LD analysis reveals five SNP pairs located at C22736T/G74357A, G34305A/G148421A, G34305A/G189246A, G148421A/G189246A, G186153A/C188379T (8 cases) with the high log of odds (>10) and strong evidence of LD (χ2 test, *P*<0.0001), in which the upper 95% confidence bound of *D*’ is above 0.98 and the lower bound is below 0.7 (Figure 2B and 2C). C22736T/G74357A SNPs are present in 28 cases, including 25 in Germany, one in Austria, one in the UK and one in Portugal. G186153A/C188379T SNP pairs has 8 German cases. There are 14 Canadian cases containing G34305A/G148421A/G189246A SNPs (Figure S1). Our results suggest that virus has evolved into at least three new lineages.

**Figure 2.**
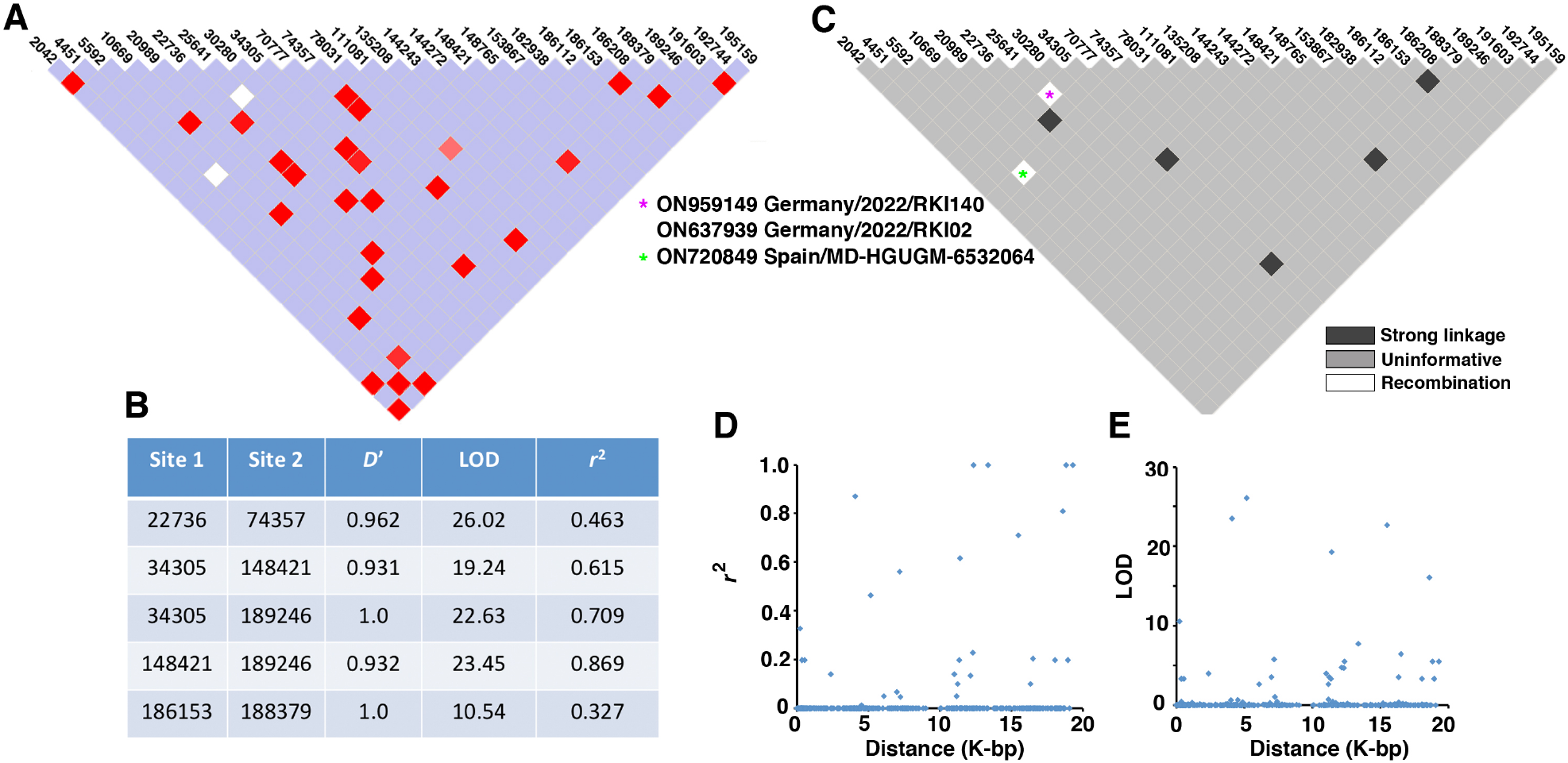
Linkage disequilibrium (LD) between SNPs in MPXV. (A) LD plot of any two SNP pairs among the 28 sites that have minor alleles in at least two strains. Each box represents a pair of SNPs. The number at the top shows the coordinate of sites in the monkeypox genome. Color in the square is given by standard (*D*’/LOD). *D*’, normalized the coefficient of linkage disequilibrium. LOD, the log of the odds. Color scheme: bright red: *D*’ = 1, LOD ≥ 2; blue: *D*’ = 1, LOD < 2, shades of pink/red: *D*’ < 1, LOD ≥ 2; white: *D*’ < 1, LOD < 2. (B) Summary of MPXV SNP pairs with strong LD (χ^2^ test, *P*< 0.001). *r*^2^, squared coefficient of correlation. (C) Haplotype blocks organization of HaploView to display the confidence bounds color scheme in (A). Strong evidence of recombination is defined as pairs for which the upper confidence bound of the *D*’ is less than 0.9 (white squares). Pairs are in strong LD if the upper 95% confidence bound of the *D*’ is above 0.98 and the lower bound is below 0.7 (black squares). (D) The *r*^2^ of each pair of SNPs (y-axis) against the genomic distance between that pair (x-axis). (E) The LOD of each pair of SNPs (y-axis) against the genomic distance between that pair (x-axis).

Moreover, the upper 95% confidence bound of *D*’ for SNP pairs C25641T/C70777T and G5592A/G78031A was 0.34 and 0.87, respectively, showing strong evidence of recombination. This result suggests that two Germany cases (ON959149 and ON637939) and one Spain case (ON720849) already gained their mutations via recombination (Figure 2B).

## Discussion

In this study, we show the first report of natural recombination of MPXV genome based on: TR (SNP-independent analysis) and LD (SNP-dependent analysis). Kugelman et al. has investigated MPXV genome diversity from 60 human samples collected in Democratic Republic of the Congo from 2005 through 2007 based on 4 regions with TRs (Kugelman et al., 2014). MPXV populations in 2022 pandemics can be categorized into 4 lineages with 11 different subgroups in clade B.1. Like Kugelman et al.’s studies, we found that none of the TR of MPXV strains were located at the protein coding region. However, all of their 60 samples (2005-2007) had identical right and left TRs. In contrast, we have detected 21 genomes (5.1%) with mismatch TR A/E during the 2022 pandemic to date (Figure 1C).

LD analysis also detected three new lineages (G22736T/G74357A, G186153A/G188379T, G34305A/G148421A/G189246A), suggesting that monkeypox virus has diverged during the 2022 pandemic. Based on neutrality test, directional selection appears to not yet be significant (normalized Wu and Fay’s *DH*= -0.74, *p* > 0.05), consistent with the idea that SNVs’ mutation rates are generally slower than recombination in poxvirus evolution (Coulson and Upton, 2011).

It has been shown TRs with the diverse length (54, 70, 125 bp) near the ends of vaccinia virus genome can provide a novel marker to detect unequal crossing over and compare the relatedness of poxviruses. (Baroudy and Moss, 1982). However, the TR diversity has been underappreciated in poxvirus study. To prevent the power of TR annotation being compromised by sequencing-assembly error, we also validated TR A to F using multiple TR detection algorithms combined with evolutionary and statistical analyses (Supplemental Materials). We have not detected ambiguous sequences at the boundary of TRs in MPXV recombinants, therefore it is very unlikely that their sequences resulted from mixed infection of different MPXV isolates. Taken together, these data confirmed that recombination did occur in 2022 monkeypox outbreak. Our results indicate that TR analysis is well-suited for detecting poxvirus recombination events. These data also demonstrated that TR and LD analysis can detect different recombination events (Figure 1F versus 2C). In combination with genomic surveillance, TR and LD analysis are both useful tools to monitor and track phylogenetic dynamics and genetic epidemiology of monkeypox transmission.

We speculate that the MPXV genomes in the 2022 outbreak, emerged most likely from a single origin, gained mutations or TRs and then evolved into different lineages and subgroups. Then, co-infection of viruses from two parental lineages occurred, followed by homologous recombination via multiple possible mechanisms (reviewed by Evans, 2022). Therefore, the progeny MPXV recombinants have mosaic pattern of TRs or mutations. So far, we have not detected any defective MPXV virus arising from a single infection.

TR insertions in the promoter and 3’-untranslated regions of MPXV may also have influence on gene expression and regulation (Supplementary Materials, Figure S2-S5). It has been reported that the 3’-to-5’ exonuclease activity of viral DNA polymerase plays an essential role in promoting extraordinarily high levels of genetic recombination in vaccinia infection (Gammon and Evans, 2009). Recombination is involved in vaccinia virus adaptation to counteract the interferon-induced antiviral host cell response mediated by the double-stranded RNA-dependent protein kinase (PKR). One of the weak PKR inhibitors, vaccinia virus K3L, undergo recurrent gene amplification with a beneficial SNV (His47Arg) via active recombination driven by selection (Elde, et al., 2012; Sasani et al., 2018). This event is mediated by vaccinia RNA polymerase and leads to rapid homogenization of K3L gene arrays (Cone et al., 2017). It is worth noting that natural MPXV C3L (vaccinia K3L homologue) is truncated at amino acid 43 by the stop codon and loss of binding site to PKRs and eIF2B (Figure S5). It is unclear whether MPXV virulence is changed due to the loss of the anti-interferon activity.

## Materials and methods

### Sequence information

Genomic sequences of MPXV (B.1 clade) from January 1 to July 20, 2022 (*N*=415) were obtained from NCBI database and available on Figshare (DOI: 10.6084/m9.figshare.20486313). FASTA files of viral sequences were downloaded and analyzed by Tandem Repeat Finder Version 4.09 (Benson, 1999). A statistical significance measurement for TRs was performed by using a cut-off alignment score [(2×match%-7×mismatch%-7×indel%)X100] by comparing them to random sequences created by simulation. (Benson, 1999). Only scores of more than 100 by Tandem Repeat Finder were considered TRs. Our TR analysis did not include small TRs (<6 bp, microsatellites), because microsatellites comprise 24% of the nucleotide sequence of these poxvirus genomes on average (Hatcher et al., 2015). The annotated TRs were validated as described in Supplemental Materials (Figure S6) and are available in Figshare (DOI:10.6084/m9.figshare.21387525). Neutrality tests and phylogenetic analysis are carried out as described in Supplementary Materials. The MPXV sequence information is summarized in Supplemental Table S1. TRs mapped on a phylogeny tree with geography information of MPXV samples are shown in Supplemental File 1 and 2.

### The linkage disequilibrium (LD) analysis

We first searched the loci via LD analysis of DNASP v6, which used the Bonferroni correction for the χ2 test to determine if the obtained LD between two loci was statistically significant (*P*< 0.001). Nucleotide polymorphism at 28 SNPs in MPXV sequences was converted by SNP_tools plug-in in Microsoft Excel to create the input files. HaploView software, version 4.1 (Broad Institute, Cambridge, USA) was used to measure and plot the normalized values (*D*’) of the coefficient of linkage disequilibrium (*D*) (Barret et al., 2005). *D*’ is defined by dividing *D* with *D*^max^, where *D*_max_ is the theoretical maximum difference between the observed and expected haplotype frequencies. HaploView was also used to calculate the squared coefficient of correlation (*r*^2^) and the log of the odds (LOD) of there being a disequilibrium between two loci. To detect recombination, we used HaploView to plot 95% confidence bounds for *D*’. Pairs are in strong LD if the upper 95% confidence bound for *D*’ is above 0.98 and the lower bound is below 0.7 (no recombination). Strong evidence for recombination is defined if pairs for which the upper confidence bound of *D*’ is less than 0.9.

## Data Availability

All sequence data are available at Figshare website: DOI: 10.6084/m9.figshare.20486313. TRAL analysis is available at Figshare website
DOI: 10.6084/m9.figshare.21387525

https://doi.org/10.6084/m9.figshare.20486313

https://doi.org/10.6084/m9.figshare.21387525

## Footnote

The methods of neutrality tests, discussion of potential influences of TRs on monkeypox genes, and supplemental figures are available in Supplemental Materials.

## Ethical approval

None declared.

## Acknowledgement

We sincerely thank scientists worldwide for providing sequence information during 2022 monkeypox outbreak. This paper is made possible by their collective work on MPXV genomic surveillance.

## Supplemental Materials

### Materials and Methods

#### Neutrality tests

Sequences are aligned using MAFFT 7 software (Katoh and Standley, 2013; Kuraku et al, 2013) and converted by MEGA 7 (Kuma et al., 2016). Neutrality tests within MPXV populations were analyzed using the polymorphism of the full MPXV genome based on the site-frequency spectrum: (1) Zeng’s *E* test, (2) Achaz’s *Y*, (3) normalized Wu and Fay’s *DH* test, and using DNASP v6 software with the camelpox virus (MZ300860) as the outgroup sequence (Rozas, et al., 2017). *DH* test is sensitive to the changes in high-frequency SNPs and primarily affected by directional selection and no other driving forces, such as demographic expansion (Zeng et al., 2006). Zeng’s *E* test contrasts high and low frequency regions of frequency spectrum, so it can detect demographic expansion after a sweep (Zeng et al., 2006). To avoid sequence errors that could reject neutrality, Achaz’s *Y* test employs the SNP number and frequency spectrum but ignores singletons, which are the class of polymorphisms that have the highest impact on several neutrality tests (such as Tajima’s *D*), (Achaz, 2008; Achaz, 2009). Recombination and the linkage disequilibrium analysis were carried out by DNASP v6 (Hudson and Kaplan, 1985; Rozas et al., 2001). Statistical significance of observed values of all tests was obtained by coalescent simulation with free recombination after 1,000 replicates. The probability for each test statistic was calculated as the frequency of replicates with a value lower than the observed statistic (two-tailed test) by DNASP v6 (Rozas, et al., 2017).

#### Phylogenetic tree

FASTA files of MPXV sequences were first aligned using MAFFT 7 software (Katoh and Standley, 2013, Kuraku et al, 2013). The neighbour-joining method and Jukes-Cantor substitution model were used to analyze phylogenetic relationships between MPXV genomes using with bootstrap resampling number set as five. The rectangular phylogenetic tree was generated by exporting the tree file in Newick format by MAFFT (Supplemental File 1). The FigTree software (version 1.4.4) was used to display the cladogram (Rambaut, 2021) after rooting with an outgroup virus sequence of the camelpox virus (MZ300860).

#### Statistical approaches for TR significance testing

We used Tandem Repeat Annotation Library (TRAL 2.0), an open source Python 3 algorithm, to identify TR seed motifs from sequence profile databases via seven *de novo* TR algorithms (HHrepID, TRED, T-REKS, TRUST, XSTREAM, PHOBOS, TRF) (Schaper et al., 2015; Delucchi et al., 2021). Briefly, circular profile hidden Markov models (cpHMMs) were built from these TR seeds and then utilized to annotate TR regions in MPXV sequences. The mutation process was described by Markov models of substitution based on the Kimura’s two parameter (K2P) model to calculate the divergence value (Kimura, 1980). All annotated TRs were statistically validated using a likelihood ratio test (LRT) that contrasts the model for the evolutionary origin of putative TR units to the scenario where the putative TRs were observed by random chance (Schaper et al., 2012). The log-likelihood for a putative TR with *n* repeat units of length *l* (ln*L*_1_), and the log-likelihood of a putative random TR under null model (ln*L*_0_) can be calculated using the equation (3) and (4) described previously (Schaper et al., 2012).

Under each of the nested models, maximized log-likelihoods can be used to construct the LRT statistic, **2(ln*L***_**1**_**-ln*L***_**0**_), which distribution was established empirically by Monte Carlo simulations to test the statistics significance (*p*-value). Each test set consisted of 1,000 simulated TRs. Gaps were treated as ambiguity characters.

## Results and Discussions

### Potential influences of TRs on MPXV genes

TR C is located at the intergenic region between A47R (IL-1/Toll-like receptor signaling inhibitor) and A49R (thymidylate kinase) (Figure S2A). Both A47R and A49R are the earliest expressed genes after virus infection (Assarsson et al., 2008; Yang et al, 2015). Vaccinia V46R (monkeypox A47R homologue) is one of determinant of viral virulence. (Stack, et al., 2005) Vaccinia mRNAs of early genes terminate within around 50 nucleotides after a UUUUUNU sequence and are subsequently polyadenylated (Yuen and Moss, 1987). Insertion of TR C is located at 14 bp downstream of the stop codon of monkeypox A47R (Figure S2B). Since TR C is also 4 bp downstream of 3’ termination sequence (UUUUUAU) of monkeypox virus, this suggests that A47R transcripts may be variable in the 3’-untranslated region between TRN=9.7 versus TRN=7.7. Whether or not the properties (degradation, location et al.) of A47R mRNA transcripts are altered require further investigation.

TR B contains a 19 bp AT-rich sequence (5’-TATATTTTATATTTTATTT-3’), and its TRN is 3.4 in 60.7% of total cases. Since sequencing information near 39.3% of TR B insertion sites is ambiguous, TR B is not included in our analysis. TR B is located at the intergenic region between A41L (chemokine-binding protein) and A42R (profilin-like protein) (Figure S3A). Since A41L and A42R transcribe in the opposite direction, this intergenic region is very likely as the promoter of A41L and A42R gene. A42R promoter contains a 5′-poly(A) leader with TAAAT and AAAAAA element, the feature of intermediate gene promoter (Yang et al., 2011; Yang et al., 2012; Yang et al., 2013) (Figure S3C). TR B is inserted at -77 bp upstream of the start codon of A42R gene, and this TR is not present in vaccinia virus genome (Figure S3B).

TR D is located at -28 bp upstream of the start codon of B19R (IFNa/b receptor glycoprotein) in the promoter region, and its copy numbers are variable (Figure S4A). Poor alignment indicates that the intergenic B18R/B19R region is very different between vaccinia and monkeypox (Figure S3B). Vaccinia B19R also has both TAAAT (−27 to -23) and AAAAAAA (−41 to -35) motif in its promoter (Yang et al., 2012), but these two elements appear to be absent in monkeypox B19R promoter (Figure S3B). Monkeypox virus has 85 bp sequence upstream of the start codon of B19R, and this 85 bp sequence does not exist in vaccinia (Figure S4C and S4D). These results suggest that gene expression of monkeypox B19R may be regulated differently from its vaccinia homologue (Figure S4B). It has been reported that the upstream core elements of both poxvirus intermediate and late promoters are clearly AT-rich sequences and regulated by TATA-binding proteins (Knutson et al., 2006). We speculate that insertion of TATA-like sequences like TR B or D may also influence gene expression of A42R and B19R.

### Phylogeny-aware and statistical approaches to validate annotated TRs

We identified TR A to F using Tandem repeat finder (TRF), which is one of the most popular TR detection algorithms (> 6,700 citations to date) (Benson et a., 1999). The TR heterogeneity has been known to contribute to the variability among different TR detection methods (Anisimova, et al., 2015). It has been aware of problems associated with TR sequences that originate from different stages during the sequencing-assembly-annotation-deposition workflow (Tørresen, et al., 2019). Because the sequencing error and gap misplacement could cause inferior quality of the predicted TR alignment, it has been shown that the TR prediction quality can be significantly improved by a phylogeny-aware gap placement method. This method appears to be promising to prevent errors in sequence alignment and evolutionary analysis (Löytynoja and Goldman 2008; Schaper et al., 2012).

To improve the accuracy and power of TR annotation, it has been suggested to apply a statistical framework in combination with a meta-approach that utilizes multiple TR prediction methods (Anisimova et al., 2015). We used Tandem Repeat Annotation Library (TRAL), which allows for evolutionary and statistics analyses to validate our annotated TRs (Schaper et al., 2015; Delucchi et al., 2021). Since MPXV genomes during 2022 outbreak most likely has a single origin (Isidro, et al., 2022), TRs in this study can be defined as related by a common ancestral unit under a Markov substitution model and a standard phylogeny model that reflects the unit duplication history (Schaper et al., 2012).

The TRAL analysis results of individual MPXV sequences of 2022 outbreak (*N*=415) are available in Figshare (DOI:10.6084/m9.figshare.21387525). After filtering the false positives with LRT statistics, TR A, C, D, E and F can be validated by the TRAL analysis with a very high statistics significance (*p* <10^−4^). TR B is omitted from the TRAL analysis, because its sequence only contains AT nucleotides (Figure 1B), which can not pass the test by the K2P model (Kimura, 1980). In addition, TRs A, D, E have very low K2P divergence values within a TR region (mean divergence value of total TRs ranged from 0.2248 to 0.2250) (Figure S6A), suggesting a consequence for TR unit gains/losses (Schaper et al., 2012). Figure S6B shows that TRAL measurement of TRNs of TR A, C, D, E and F, which recombination patterns are consistent with Figure 1F. Taken together, our data confirmed that MPXV recombination can be detected by multiple TR analyses, and it is not affected by the sequencing-assembly error.

### The potential roles of APOBEC3 in monkeypox virus mutations

Apolipoprotein B mRNA editing catalytic polypeptide-like3 (APOBEC3) is not degraded by vaccinia virus infection, and it is also not required for vaccinia virus replication (Kremer, et al., 2006). However, it has been suggested that host APOBEC3 in viral evolution with signs of potential human adaptation of MPXV in ongoing microevolution (Isidro et al., 2022). Gigante et al. reported that APOBEC3 activity has been recurrent and dominant in West African MPXV evolution (2017-2022) recently (Gigante et al., 2022). When compared with previous MPXV sequences, many mutations in the 2022 MPXV sequences were 5’ GA-to-AA, suggesting APOBEC3 activity (O’Toole and Rambaut, 2022). 91% G-to-A mutations were present in an APOBEC3 motif (160 out of 176). Most APOBEC3 G-to-A mutations were specifically GA-to-AA (156 out of 160, 96%). These results suggested that other APOBEC3 subfamily members, but not APOBEC3G (GG-to-AG changes), are involved in the editing. So far, we have identified neither GA-to-AA nor GG-to-AG changes in TR regions based on Isidro et al.’s <u>Python script</u> (Isidro et al., 2022). It will be interesting to explore APOBEC3 roles in MPXV recombination in the future.

---

Supplemental File 1. This file contains the Newick tree of the MPXV sequence dataset for Supplemental File 2.

Supplemental File 2. Rooted phylogenetic tree of MPXV genomes during 2022 pandemics. Alignments of viral sequences were generated using MAFFT, and the phylogenetic tree was visualized using FigTree. Rooting was done by introducing the camelpox virus sequence (MZ300860) as an outgroup virus. Group U, M, and I of MPXV genomes is labeled in green, blue, and red, respectively. Recombinant MPXV isolates based on TR analysis are labeled in orange.

Supplemental Table 1. Summary of MPXV genome sequence information in this study.

**Figure S1.**
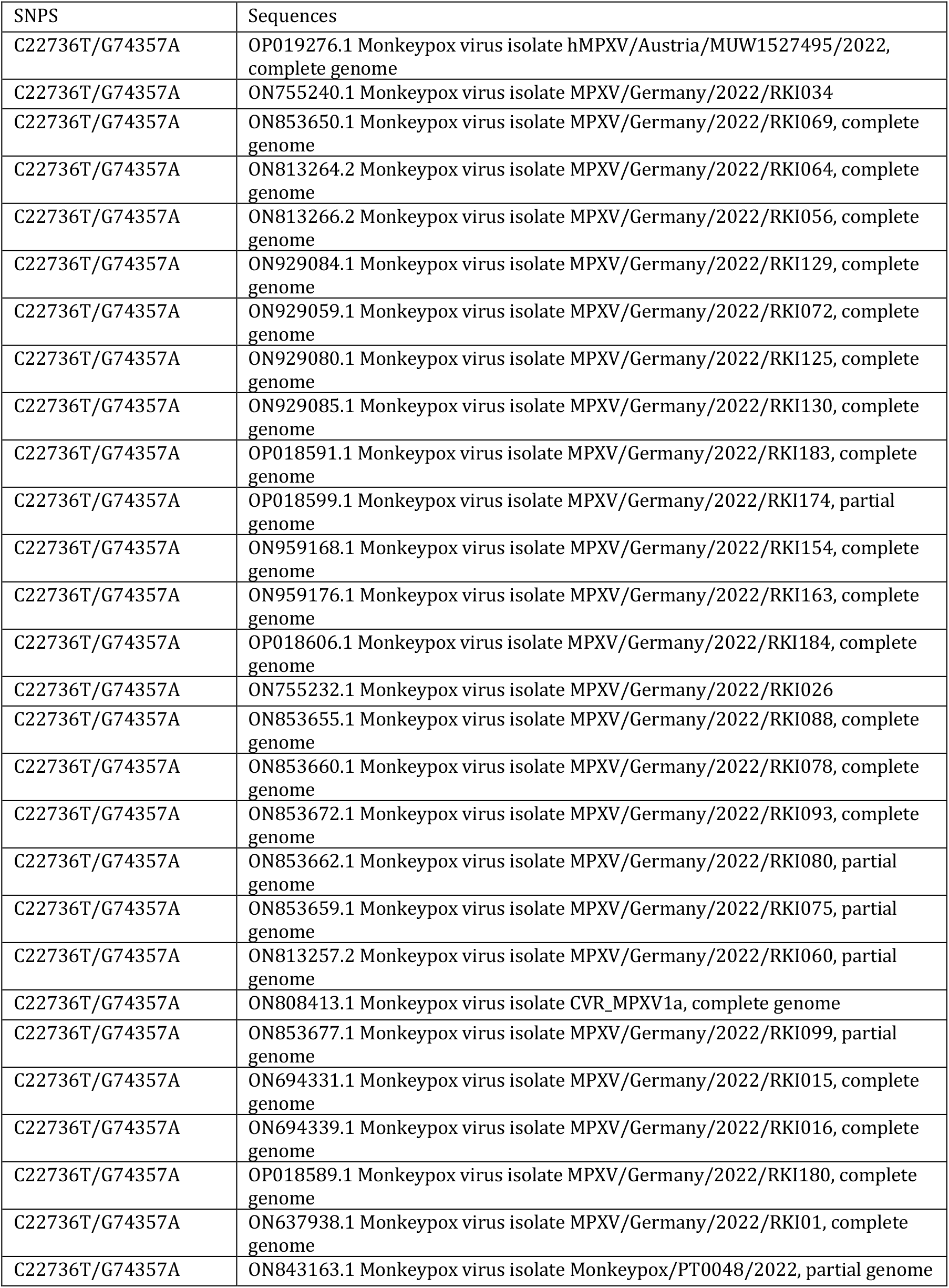
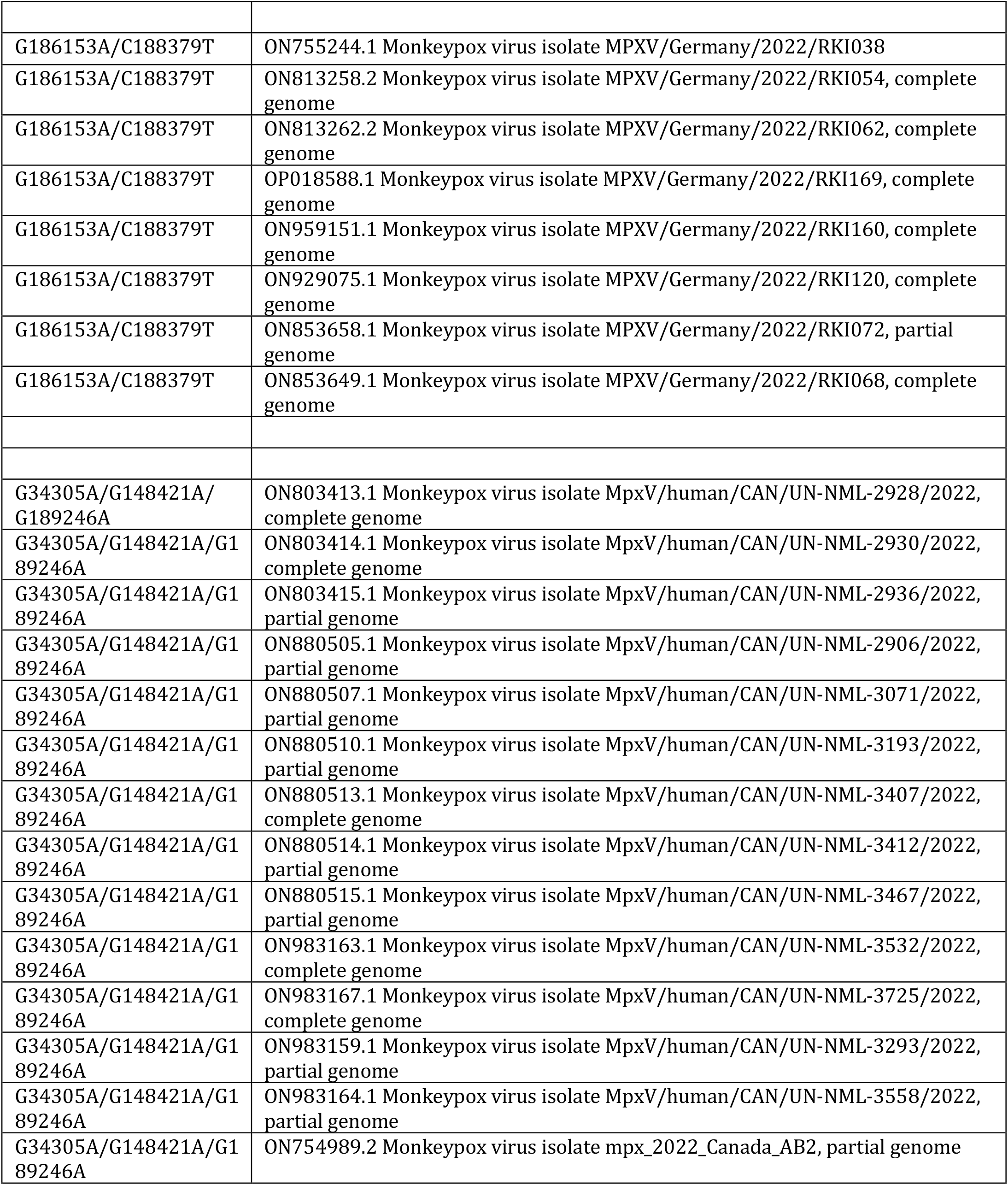
Sequences that contain SNP pairs with strong linkage disequilibrium, which the upper 95% confidence bound of the *D*’ is above 0.98 and the lower bound is above 0.7.

**Figure S2.**
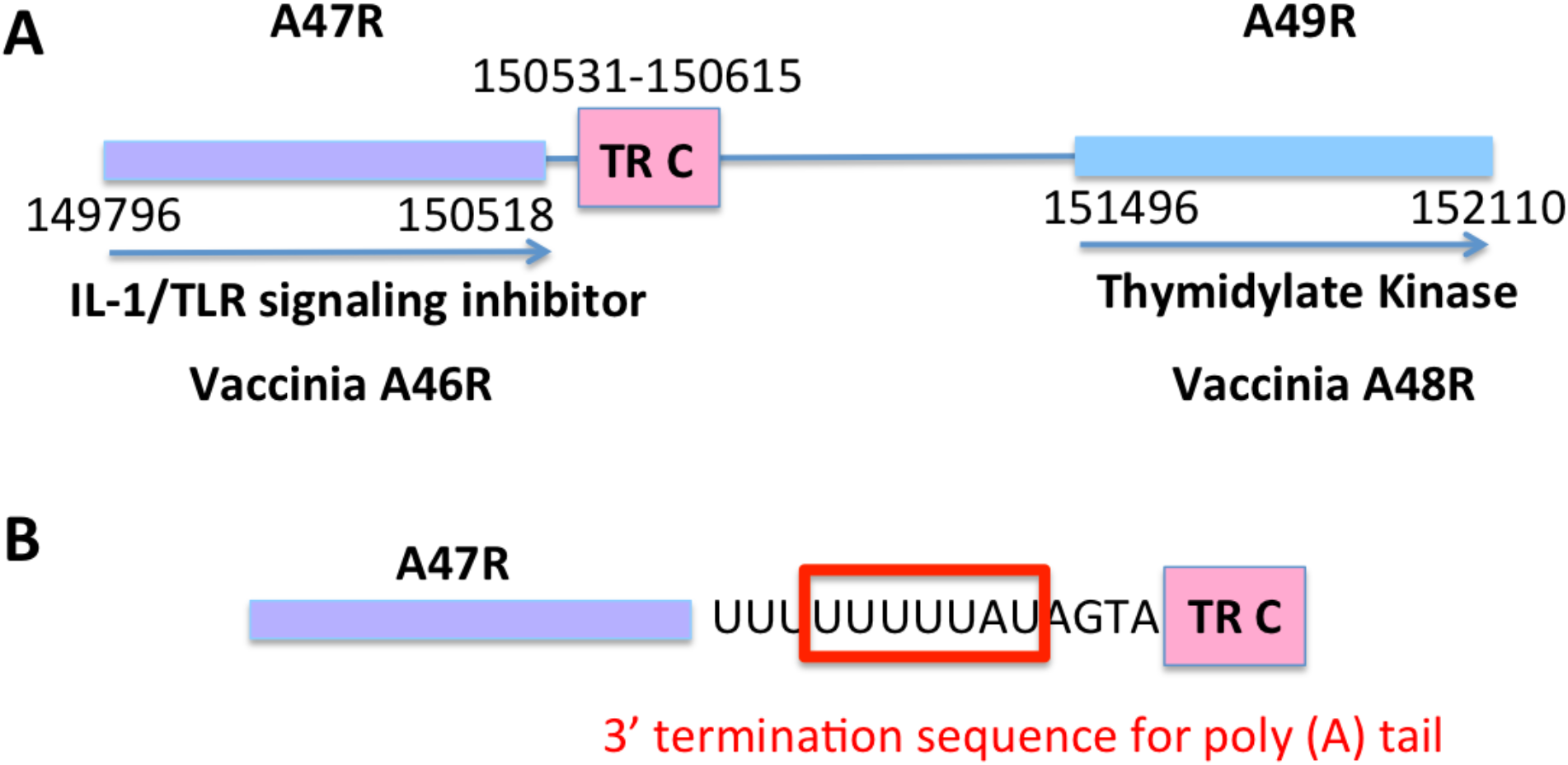
TR C in the intergenic region of monkeypox A47R and A49R. (A) The map of TR C, A47R, and A49R gene. Arrows indicate the gene direction. (B) The location of TR C at the 3’-untranslated region of A47R transcript. The 3’ termination UUUUUNU sequence for adding poly(A) tail is boxed in red. Reference MPXV sequence is OP19276.

**Figure S3.**
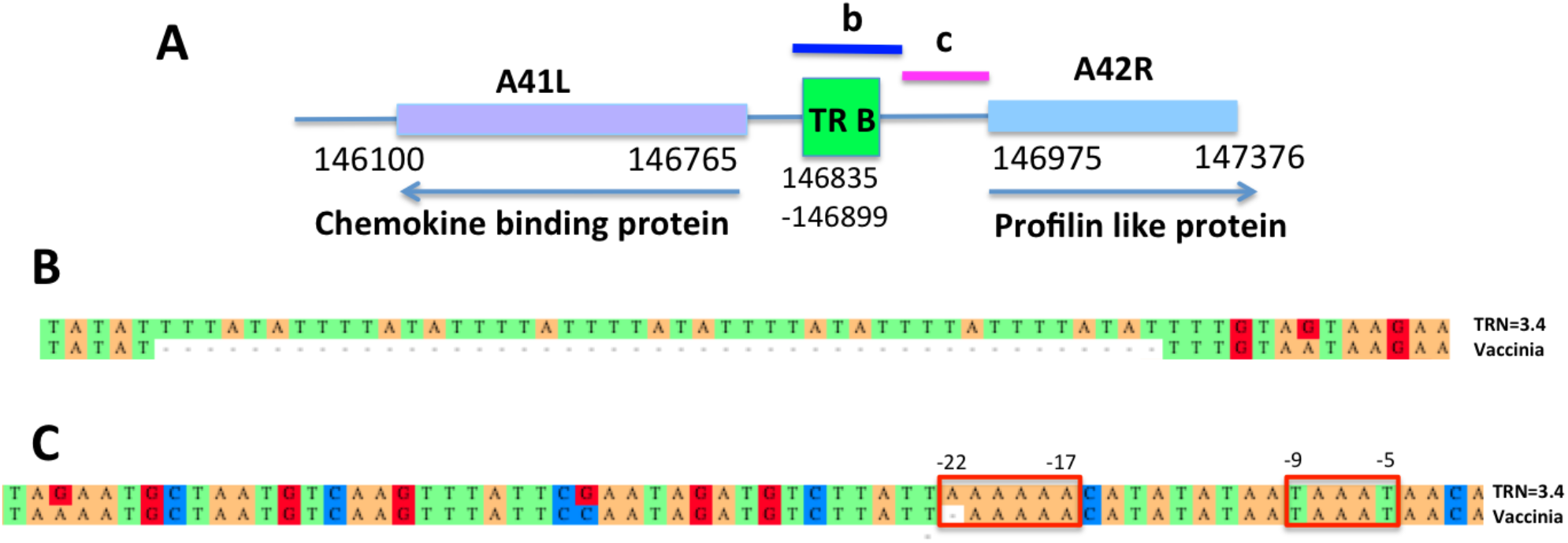
TR B in the intergenic region of monkeypox A41L and A42R. (A) The map of TR B, A41L, and A42R gene. Arrows indicate the gene direction. Sequence b and c are shown in (B) and (C). (B) Sequence of TR B insertion in monkeypox genome as in (A) compared to vaccinia sequence. (C) Alignment of monkeypox and vaccinia in A42R promoter. TAAAT and AAAAAA element are boxed in red. Reference MPXV sequence is OP19276.

**Figure S4.**
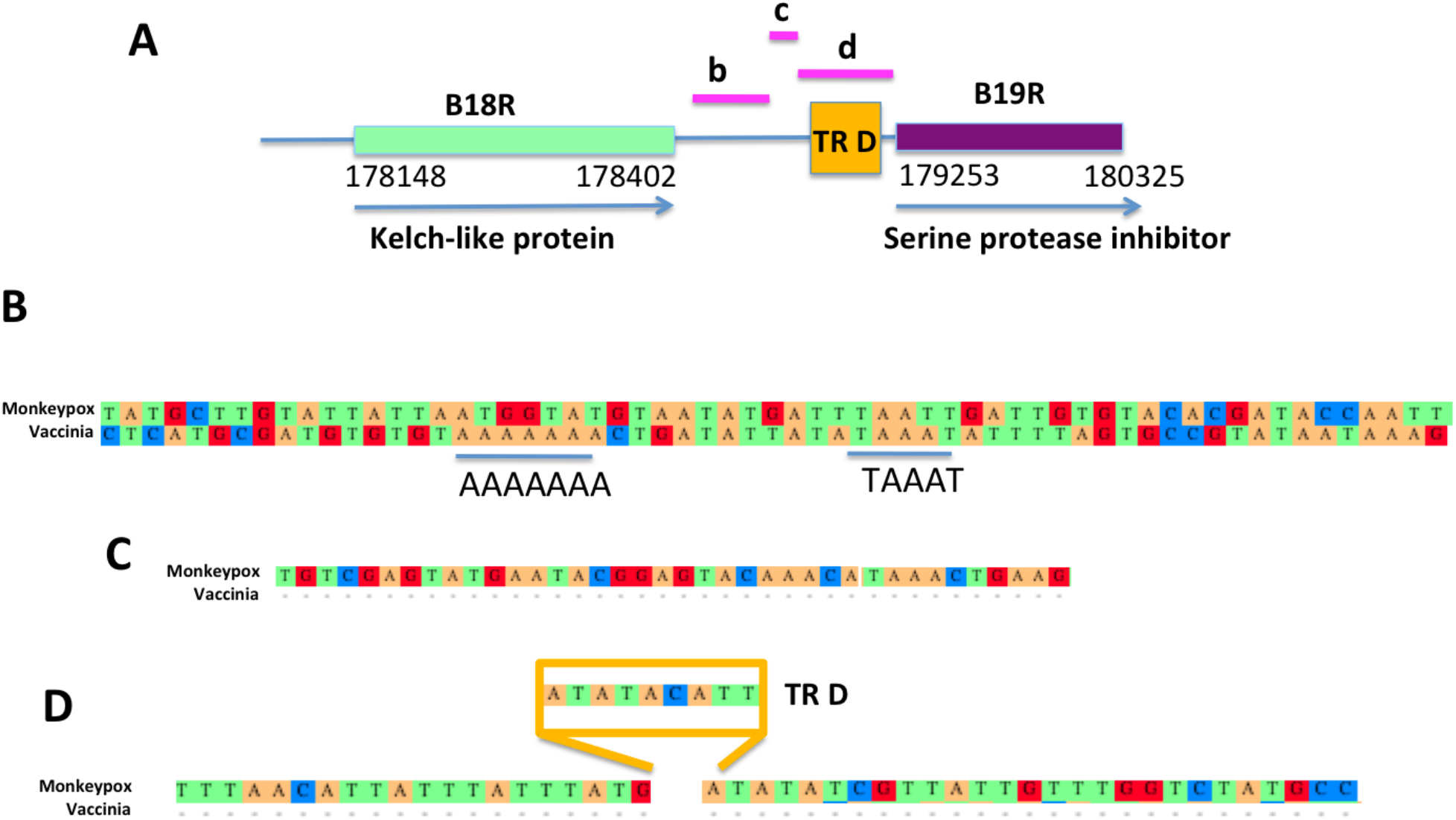
TR D in the intergenic region of monkeypox B18R and B19R. (A) (A) The map of TR D, B18R, and B19R gene. Arrows indicate the gene direction. Sequence b, c and d are shown in (B), (C) and (D). (B) Alignment of monkeypox sequence B with vaccinia B19R promoter, including TAAAT and AAAAAAA element only found in vaccinia. (C and D) Sequences of B19R promoter of monkeypox. Sequence of TR D insertion in monkeypox genome as in (A). Reference MPXV sequence is OP19276.

**Figure S5.**
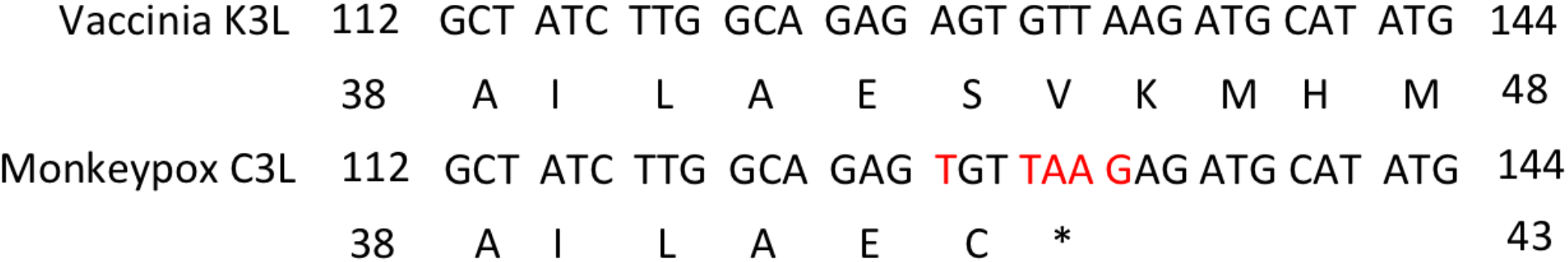
Comparison of vaccinia K3L and monkeypox C3L sequence. The stop codon was introduced at bp 130-132, causing C3L a truncated protein compared to K3L.

**Figure S6.**
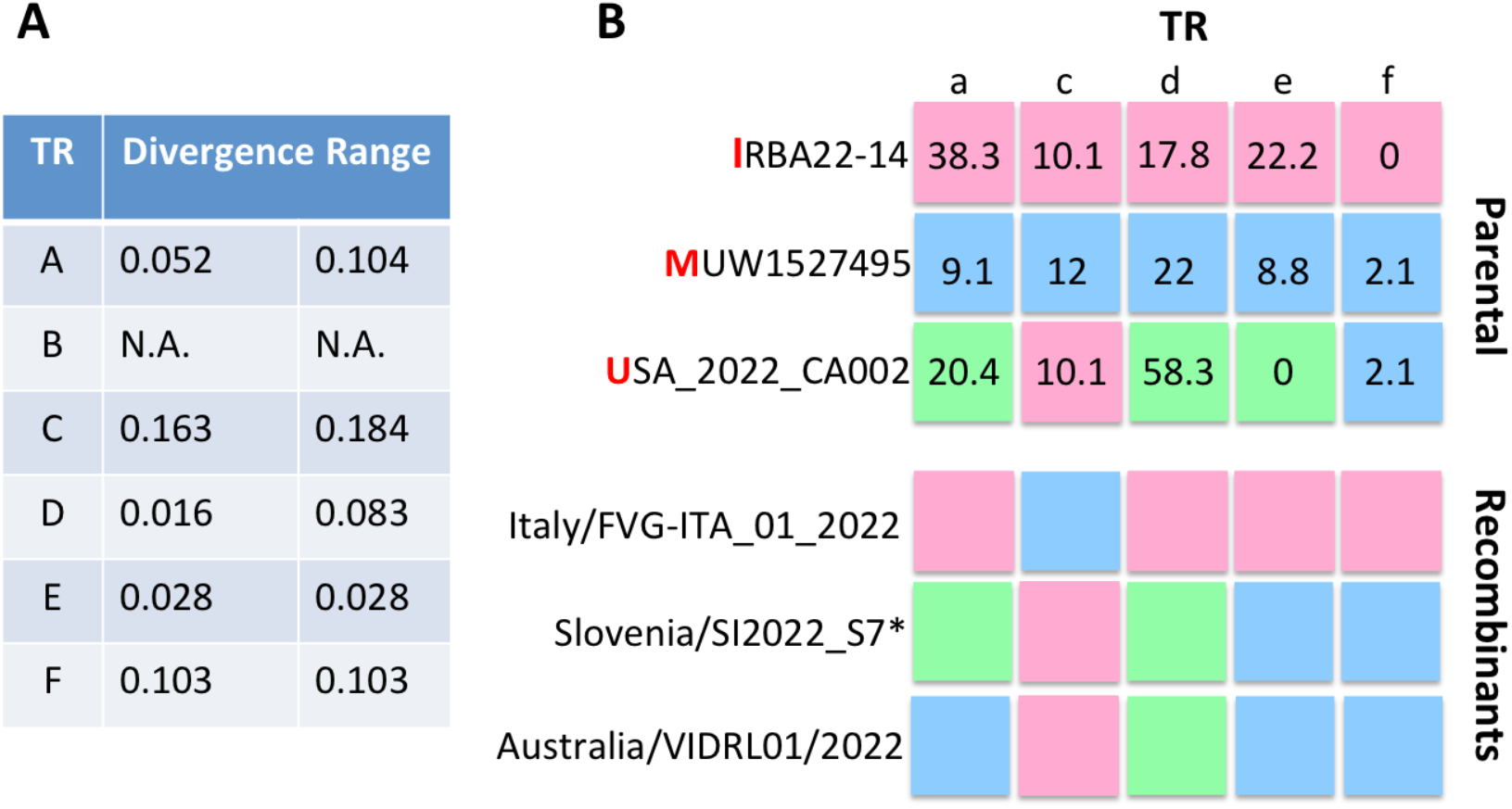
(A) Table of divergence value ranges of TR A to F. The value of TR B is not available because its sequence is AT only. (B) Each box (TR A, C, D, E, F) contains individual TRNs with different colors associated with the parental viral sequences: IRBA22-14 (I, ON755040), MUW1527495 (M, ON019276) or USA_2022_CA002 (U, ON954773) as Figure 1F. TRs and TRNs were validated by the TRAL algorithm with *p*-value < 10^−4^.

