## Supplementary figures and images for "Recombination shapes 2022 monkeypox outbreak"

### Suppemental File 2.pdf

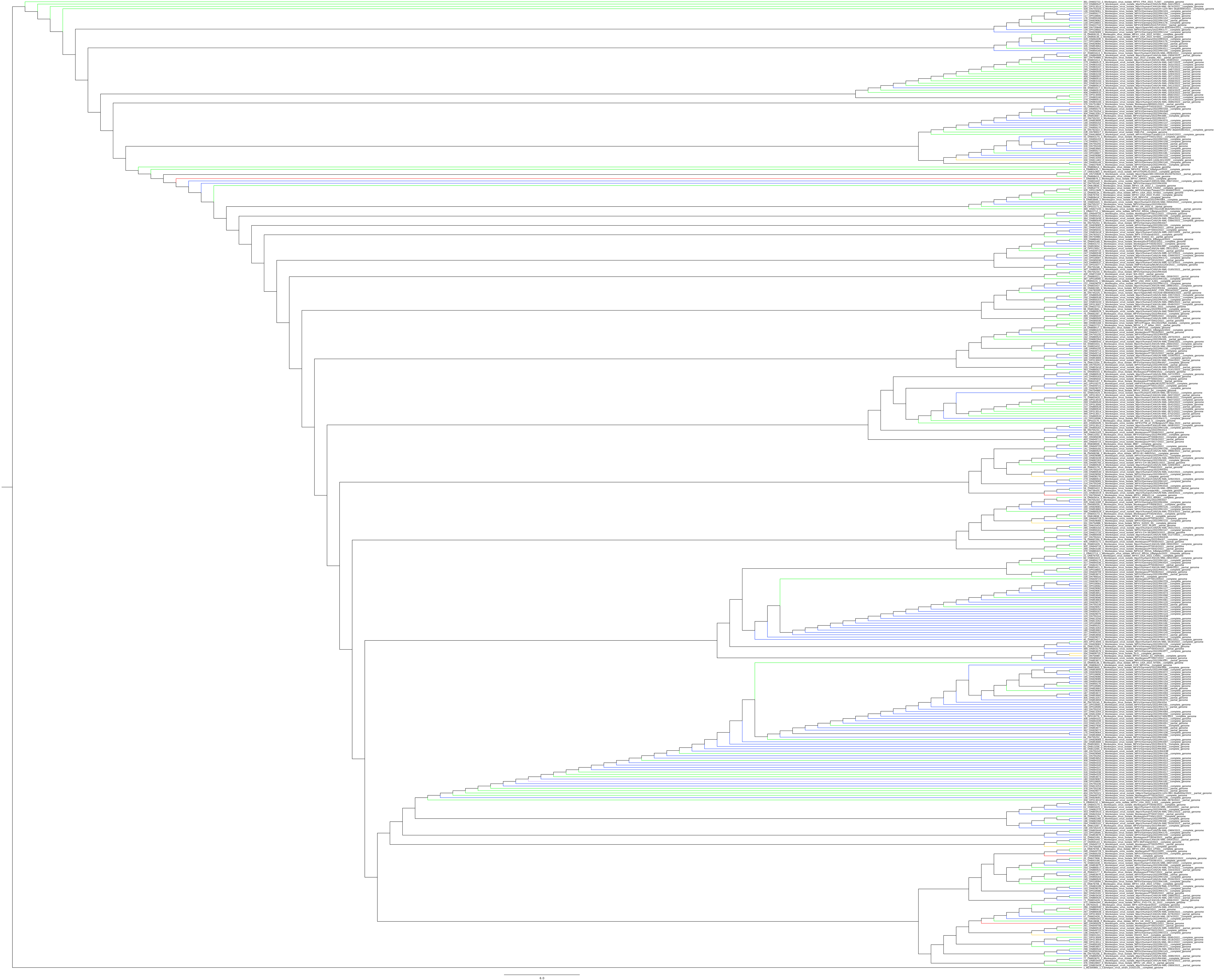
